# The development and usability testing of two arts-based knowledge translation tools for pediatric procedural pain

**DOI:** 10.1101/2021.06.17.21259118

**Authors:** Anne Le, Lisa Hartling, Shannon D. Scott

## Abstract

Procedures carried out in acute care settings, such as emergency departments (EDs), are among the most common sources of acute pain experienced by children. Such procedures may include intravenous insertions (IVs), venipuncture, and wound irrigation and repair. Inadequately managed procedural pain can cause negative short-term and long-term implications for children, ranging from anxiety to aversion to healthcare. Parents have repeatedly expressed that they do not have the necessary tools to comfort or distract their child during uncomfortable medical procedures. As such, the purpose of this study was to work with parents to develop and evaluate two digital tools for pediatric procedural pain.

A whiteboard animation video and interactive infographic were developed following a systematic review and interview with parents. Prototypes were tested in five ED waiting rooms in two Canadian provinces. Sites included those in urban, rural, and remote settings. Overall, parents rated the tools highly, suggesting that engaging with parents to develop arts-based digital tools is a highly effective method in ensuring that parents can understand and utilize complex health information.

**Author Contributions:** This study was conducted under the supervision of Drs. Shannon D. Scott (SDS) and Lisa Hartling (LH), PIs for **translation Evidence in Child Health to enhance Outcomes** (ECHO) Research and the **Alberta Research Centre for Health Evidence** (ARCHE), respectively. Both PIs designed the research study and obtained research funding through Translating Emergency Knowledge for Kids (TREKK) Networks of Centres of Excellence of Canada (NCE).

SDS designed and supervised all aspects of tool development and evaluation.

LH co-designed and supervised the qualitative study involving interviews with parents and systematic review of parent experiences and information needs.

Tony An developed the infographic.

Kassi Shave conducted and analyzed qualitative interviews with parents.

Anne Le (AL) conducted usability testing.

AL analyzed usability data.

All authors contributed to the writing of this technical report and provided substantial feedback.

This work was funded by:

**Networks of Centres of Excellence:**

- Klassen, T., Hartling, L., Jabbour, M., Johnson, D., & Scott, S.D. (2015). Translating emergency knowledge for kids (TREKK). Networks of Centres of Excellence of Canada Knowledge Mobilization Renewal ($1,200,000). January 2016 – December 2019.

**Women and Children’s Health Research Institute (WCHRI):**

- Scott, S.D & Hartling L. (2016). Translating Emergency Knowledge for Kids renewal. Women and Children’s Health Research Institute (matched dollars, $150,000). April 2016 – December 2019.

**This report should be cited as:** Le, A., Hartling, L., Scott, S.D. (2021). The development and usability testing of two arts-based knowledge translation tools for pediatric procedural pain. Technical Report. ECHO Research, University of Alberta.

Available at: http://www.echokt.ca/research/technical-reports/

## Introduction

Procedures regularly carried out in acute care settings, such as emergency departments (EDs), are among the most common sources of acute pain experienced by children [1]. Poorly managed pain can cause negative short-term and long-term implications for children, ranging from anxiety to aversion to healthcare [2-6] In the ED, distress and anxiety may be heightened due to procedures typically being unplanned, leading to parents and children not feeling prepared [7]. Despite the available and widely advocated evidence-based interventions for procedural pain management, the implementation of these interventions in healthcare settings has been minimal [8-9].

Research has found that many parents are unsure of how to effectively comfort or distract their child during a medical procedure, or do not feel confident doing so [10-16]. Knowledge translation (KT) tools may empower parents to take an active role in the management of their child’s procedural pain. In particular, visually engaging media such as videos or infographics hold great potential for communicating health research in an easily accessible and understandable manner.

Translating Emergency Knowledge for Kids (TREKK) is a national program in Canada with the aims of developing knowledge translation (KT) tools that encompass the best available evidence for pediatric health care providers and parents of children with acute illnesses. In our role as co-directors of TREKK, we have overseen the development of several KT tools for parents of children with acute conditions. This aligns with our roles as principal investigators of Translating Evidence in Child Health to enhance Outcomes (ECHO) and Alberta Research Centre for Health Evidence (ARCHE) research groups at the University of Alberta.

The purpose of this study was to work with parents to develop and assess the usability of an interactive infographic and video for procedural pain in children.

## Methods

This multi-method study utilized patient engagement to develop, refine, and evaluate a whiteboard animation video and interactive infographic for pediatric procedural pain. Research ethics approval was obtained from the University of Alberta Health Research Ethics Board (Edmonton, AB) [Pro00062904]. Operational approvals were obtained from the local health authorities to conduct usability testing in their emergency department.

### Compilation of Parents’ Narratives

A project coordinator trained in qualitative methodology conducted semi-structured interviews with parents (n = 12) of children who required a procedure involving a needle at the Stollery Children’s Hospital (Edmonton, Canada). Semi-structured interviews were selected in order to cover all aspects of parents’ experience with procedural pain while simultaneously allowing parents to freely share and discuss their perspective (**Appendix A**). Interviews were recorded, transcribed verbatim, and managed using NVivo-10. Concurrently, a systematic review was conducted to review the literature on parents’ experiences and information needs related to pediatric procedural pain. Results from the qualitative study and systematic review are published elsewhere [2, 17].

### Intervention Development

Results from the systematic review were combined with qualitative data to generate themes and key quotes to be included in the content outlines for the tools. Along with best evidence from the TRanslating Emergency Knowledge for Kids (TREKK) Bottom Line Recommendations (BLR) for managing procedural pain in children, researchers then developed a video script and infographic skeleton to be shared with illustrators, video developers, and graphic designers for prototype development [18]. Information on how to manage procedural pain was embedded within the storyline of the video, which depicted parents struggling to manage their child’s pain. Similarly, information on distraction and pain management techniques was included in the infographic.

### Video Intervention

Several iterations of the procedural pain whiteboard animation video were developed before a version was finalized for usability testing. The first prototype of the whiteboard animation video was almost 4 minutes in length and included images that were ambiguous to ensure inclusivity of all ethnicities and genders. Characters were assigned a colour (i.e., mother = purple, father = blue) and the child was given a unisex name, “Sam”. Stakeholders found the colouring to be distracting and thus, colours were removed prior to usability testing.

The next iteration of the video (that underwent usability testing) was 2 minute 58 seconds in length and was predominantly in black and white (**Appendix B**). Concepts and information that were considered important were changed to red to highlight their importance. The English-language video began with a mother and a father sitting in the emergency department with their ill 6-year-old child named Sam. The narrator (third person) describes Sam as being sick and requiring a needle poke. As the video is narrated, the images on the screen change, providing accompanying illustrations and words to emphasize key points. The video provided parents with various reasons why a child would require a needle poke, pain management techniques, and ways to reduce their child’s stress during a procedure. At the end of the video, this information is reiterated.

### Infographic Intervention

As with the whiteboard animation video, the interactive infographic also underwent several rounds of development prior to usability testing. The infographic was developed to function similarly to a digital application (“App”), such as those used on mobile phones or handheld tablets (**Appendix C**). Users were directed to a “home screen”, which contained a menu, allowing them to select and view information based on their own needs. The menu items included on the home screen included, “Why does my child need a needle poke?”, Tips for Everyone, Age Specific Solutions, and Useful Links. In total, the infographic included five different pages.

### Revisions

The procedural pain tools were developed using iterative processes. Parents from our Pediatric Parental Advisory Group (PPAG), health care providers, and the study team provided continuous feedback over the course of development. These groups were asked to assess the tools for quality of information and evidence, length of the tool, aesthetics, usefulness, and perceived value. The PPAG members were asked to comment on tool relevance, length, and usefulness. Our research team also holds weekly meetings to discuss the development of our tools.

### Focus Group with Children

5 children (aged 7-14 years) were recruited through convenience and snowball sampling and interviewed within a focus group setting (**Appendix D**). The purpose of this focus group was to understand children’s experience with procedural pain that took place in a hospital setting, to help in the development of the tool. Participants were asked about the language used to describe aspects of the experience (i.e., “poke” or “needle”) and to provide detailed descriptions of their experience (reason for receiving a needle, how they felt, if any techniques to mitigate pain were used, and preferred techniques). Following these discussions, the children were asked to view the first prototype of the tool and give their feedback. See Image 1 for our KT Tool Development Process.

**Image 1.**
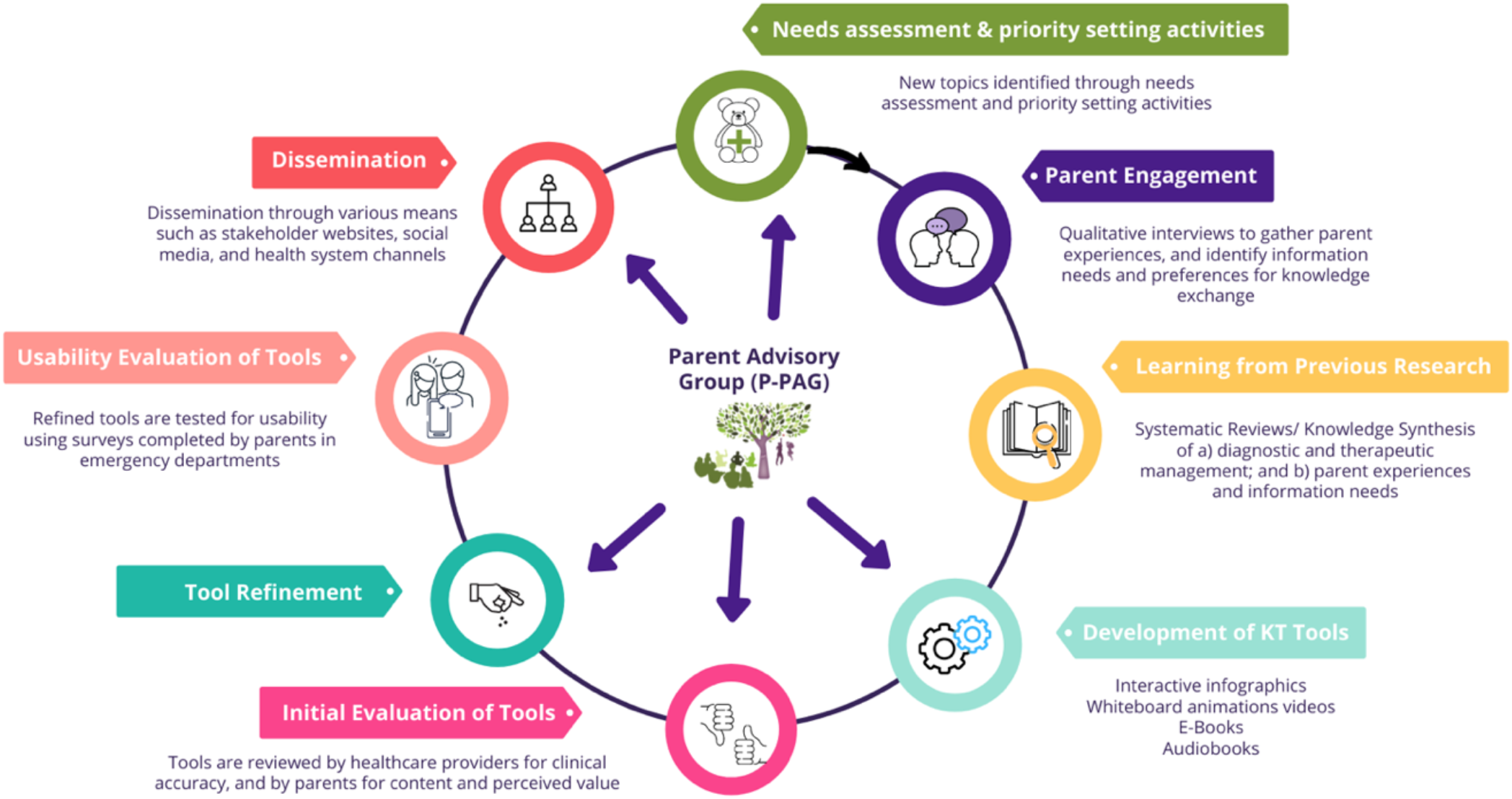
KT Tool Development Cycle

### Surveys

Survey data were collected via iPads in five ED waiting rooms in Canada, including those from remote, rural, and urban regions. These included the Cobequid Community Health Centre, Hants Community Hospital, and Colchester East Hants Health Centre in Nova Scotia, Stanton Territorial Hospital in the Northwest Territories, and Stollery Children’s Hospital in Alberta. Members of the study team approached parents in the ED to determine interest and study eligibility. Eligible and interested parents were presented with the electronic consent forms loaded onto the iPad and asked to answer the following question to give consent, “Do you consent to participate in the study?”. Participants who answered “no” would be redirected to a “thank-you” page whereas parents who answered “yes” would be redirected to the survey. Surveys were developed using SimpleSurvey, a secure, Canadian platform and stored on iPads for data collection.

Surveys were informed by a systematic review of over 180 usability evaluations and comprised of 9, 5-point Likert items assessing: 1) usefulness, 2) aesthetics, 3) length, 4) relevance, and 5) future use [19]. Likert items ranged from *strongly agree* to *strongly disagree*. Parents were also asked to provide their positive and negative opinions of the tool via two free text boxes. Study team members were available in the ED to provide technical assistance and answer questions as parents were completing the surveys (**Appendix E**).

### Data Analysis

Data were cleaned and analyzed using SPSS v.24. Descriptive statistics and measures of central tendency were generated for Likert items. Each answer on the Likert scales was given a corresponding numerical score from 5 to 1, with 5 being strongly agree 1 being strongly disagree [20-21]. T-tests were conducted to compare the two tools. Open-ended survey data were analyzed thematically. A summary of parents’ responses from the usability surveys was shared with the creative team to allow for final revisions to be made.

## Results

A total of 73 participants evaluated the e-tools. 27 participants evaluated the procedural pain video and 46 evaluated the infographic. The reason for the large discrepancy in participants between the two tools is because the infographic underwent some minor changes during usability testing to improve ease-of-use of the tool. For instance, the first iteration of the infographic did not have a clear and distinct “back” button to allow users to return to the main menu. Rather, the button was designed to be minimalistic (plain white arrow pointing in the “back” direction), which resulted in the first set of usability participants missing this completely. Users either did not further explore the tool or mistakenly selected the browser return button, which navigated them away from the infographic completely. As such, part way through usability testing, the infographic was updated to include a more noticeable and distinct back button so that participants could view and evaluate it fully.

**Table 1.** Demographic characteristics of parents who assessed the usability of the procedural pain e-tools (N=73)

| Characteristic | n (%) |
| --- | --- |
| Sex |  |
| Female | 47 (64.4) |
| Male | 26 (35.6) |
| Age |  |
| 20-29 years | 10 (13.7) |
| 30-39 years | 26 (35.6) |
| 40-49 years | 29 (39.7) |
| 50-59 years | 7 (9.6) |
| 60 years and older | 1 (1.4) |
| Marital Status |  |
| Married/Partnered | 54 (74.0) |
| Single/Separated/Divorced/Widowed | 19 (26.0) |
| Education |  |
| Some high school | 5 (6.8) |
| High school diploma | 9 (12.3) |
| Some post-secondary | 13 (17.8) |
| Post-secondary certificate/diploma | 19 (26.0) |
| Post-secondary degree | 19 (26.0) |
| Graduate degree | 6 (8.2) |
| Other | 1 (1.4) |
| Missing | 1 (1.4) |
| Household Income |  |
| Less than \$25,000 | 6 (8.2) |
| \$25,000-\$49,000 | 7 (9.6) |
| \$50,000-\$74,000 | 14 (19.2) |
| \$75,000-\$99,000 | 12 (16.4) |
| \$100,000-\$149,000 | 14 (19.2) |
| \$150,000 and over | 8 (11.0) |
| Prefer not to answer | 11 (15.1) |
| Missing | 1 (1.4) |
| Number of Children in the Family |  |
| 1 | 22 (30.1) |
| 2 | 29 (39.7) |
| 3 | 12 (16.4) |
| 4 | 6 (8.2) |
| 5+ | 3 (4.1) |

Overall, parents rated the whiteboard animation tool positively, with most parents answering *agree* or *strongly agree* on all usability questions. Furthermore, when converted to numerical values, we obtained means of at least 3.86 out of 5.00 on the usability questions. 25 selected strongly agree and agree to the question “it is useful” (mean = 4.19) and 24 selected strongly agree and agree when asked whether the tool was “simple to use” (mean = 4.19). Parents also felt that the length was appropriate, with 24 selecting strongly agree or agree (mean = 4.15). Parents also deemed the tool as relevant to them as parents (mean = 4.11) and that it could be used without additional help or instructions (mean = 4.15), with 24 selecting strongly agree or agree for both questions. When asked if parents would use the tool in the future or if it would help them make decisions about their child’s health, 23 strongly agreed or agreed (mean = 4.04) and 24 strongly agreed or agreed (mean = 4.26), respectively. Parents gave slightly lower scores when asked about the tool’s aesthetics and if they would recommend it to a friend, yielding 3.89 and 3.85, respectively.

Likewise, parents also rated the interactive infographic tool positively, with scores of at least 3.74 on all usability items. Parents found the tool useful (mean = 4.35), relevant (4.37), and simple to use (4.37). When asked whether they would be able to use the tool without written instructions, parents mainly strongly agreed or agreed, yielding a mean of 4.09. Furthermore, parents felt that the length of the infographic was appropriate (4.35) and that it was aesthetically pleasing (4.48). As a result, they were also very likely to use the tool in the future (3.82), to use it to make decisions about their child’s health (3.74), and recommend the tool to their friends (4.39).

When comparing participant responses for the video versus the infographic, scores for aesthetics and whether they would recommend the tool to a friend were significantly higher for the infographic (p < 0.05). The score for whether the tool would help parents make decision about their child’s help was significantly higher for the video.

**Table 2.** Means (SD) of participant responses to the usability survey

| Usability Measures | Video | Infographic | P value* |
| --- | --- | --- | --- |
| It is useful. | 4.19 (0.92) | 4.35 (0.71) | 0.43 |
| It provides information that is relevant to me as a parent. | 4.11 (1.05) | 4.37 (0.65) | 0.20 |
| It is simple to use. | 4.19 (0.88) | 4.37 (0.71) | 0.33 |
| I can use it without written instructions or additional help. | 4.15 (0.86) | 4.09 (0.96) | 0.79 |
| Its length is appropriate. | 4.15 (0.86) | 4.35 (0.71) | 0.28 |
| It is aesthetically pleasing (i.e. images, colours, etc.). | 3.89 (1.05) | 4.48 (0.59) | 0.003* |
| It helps me make decisions about my child's health. | 4.26 (0.66) | 3.74 (0.95) | 0.02* |
| I would use it in the future. | 4.04 (0.94) | 3.82 (1.04) | 0.39 |
| I would recommend it to a friend. | 3.85 (1.13) | 4.39 (0.65) | 0.01* |

\*p < 0.05

In open text boxes, parents described the video as “simple and clear” and “helpful”. Parents were particularly interested by suggestions to use numbing cream prior to medical procedures, stating that they did not know that this was an option. Parents were also receptive to the idea of providing their child with encouragement, one stated: “I like the idea of being honest and encouraging. Not saying sorry is an important message to parents. Their attitudes and discussions about it influence reactions” (*Participant 13*).

Comments were similar for the infographic, with parents saying that the tool would be useful for younger parents and that the design was clean.

Two issues were brought up which resulted in both the video and infographic undergoing drastic changes prior to their release. First, many parents had issues with the buttons (nonresponsive, hitting the wrong buttons, etc.) on the infographic so the tool was drastically modified to improve ease of use. In addition, parents and health care partners did not like the illustrations (in particular the characters) used in the whiteboard animation video due to their “unrealistic nature”; thus, we commissioned new illustrations for our suite of whiteboard animation tools.

### Final Video

The final video was 2 minutes 58 seconds in length and featured a new set of characters developed to look more realistic (**Appendix F**). As in the original, characters were made to look ambiguous in terms of race. However, the characters, storyline and pain management strategies remained the same.

### Final Infographic

The final infographic was changed to resemble a webpage (**Appendix G**) rather than an application. It allows users to scroll through the information and was similar to the App style in that it allowed users to explore the information at their own pace. However, rather than having parents navigate through different links, all the information now resides on the same page. The information provided in the infographic was unchanged; though, like the video, the illustrations were modified to look more realistic.

## Conclusions

The purpose of this study was to develop and evaluate an interactive infographic and whiteboard animation video for procedural pain management. Using a multi-phase method that engaged parents, children, health care professionals and researchers, we developed a tool that was highly rated amongst its intended end users: parents. Overall, responses to our usability survey were very positive with most parents agreeing or strongly agreeing with all of our usability items. This indicates that our whiteboard animation tool and interactive infographic are appropriate resources for parents with children undergoing painful procedures. These findings have several implications.

A user-centred study design that includes and engages stakeholders in the co-development of health education tools or decision-making aids is immensely valuable. Resources that are developed for end-users with the help of end-users increases the relevancy of the tool for the target population. We aimed to achieve this throughout our entire tool development and evaluation process as visualized in Image 1; first, by conducting our national needs assessment that informed us of the pediatric conditions parents wanted more information on [22], then by conducting a systematic review of the literature examining parent information needs and experiences regarding procedural pain management [17], and finally conducting further interviews with parents on this specific health condition [2]. As such, prior to generating narratives for our tool, we had a strong understanding of parent information needs and preferences. We further engaged parents by creating collaborative environments to collect feedback on our tools through our parental advisory group [23]. Finally, we collected feedback from parents in the emergency department and modified the tools based on this feedback. The result is a tool that aligns with parents’ preferences on usability, aesthetics, ease-of-use, and length. In addition to this, results for questions regarding future use and whether parents would recommend the tool to their friends were also rated highly, suggesting that both content and method of dissemination were suitable for parents.

## Data Availability

Data from this project are stored on secured servers at the University of Alberta. Due to privacy and confidentiality agreements, they are not available for viewing or use by individuals outside the study team

## Other Outputs from this Project

## Appendices

### Appendix A – Interview Guide

Parents will be interviewed to understand their experience having a child with procedural pain. Semi-structured interviews will be conducted with parents in order to get their “narrative” or experiences. The following questions will be used to guide these interviews. Being true to semi-structured interview techniques, interview questions will start broad and then move to the more specific.

1. Tell me about your experience having your child experience procedural pain.
2. Tell me about your child that was ill. How old is your child? How was your child ill? Has your child previously had procedural pain?
3. How did you feel during this experience?
4. What did you do to manage/prevent symptoms of procedural pain? (any techniques you used, for example, talking with child before or after, reading a storybook, talking with family/friends, etc.)
5. What strategies were put in place by health care professionals to help your child? (for example, using distraction, numbing medication). Did they ask you to do anything?
6. How did your child manage the experience? How did you feel about the outcome of this situation?
7. If presented with the same situation again, would you do anything differently? If so, please tell me.

### Appendix B – Images from Usability Testing Video

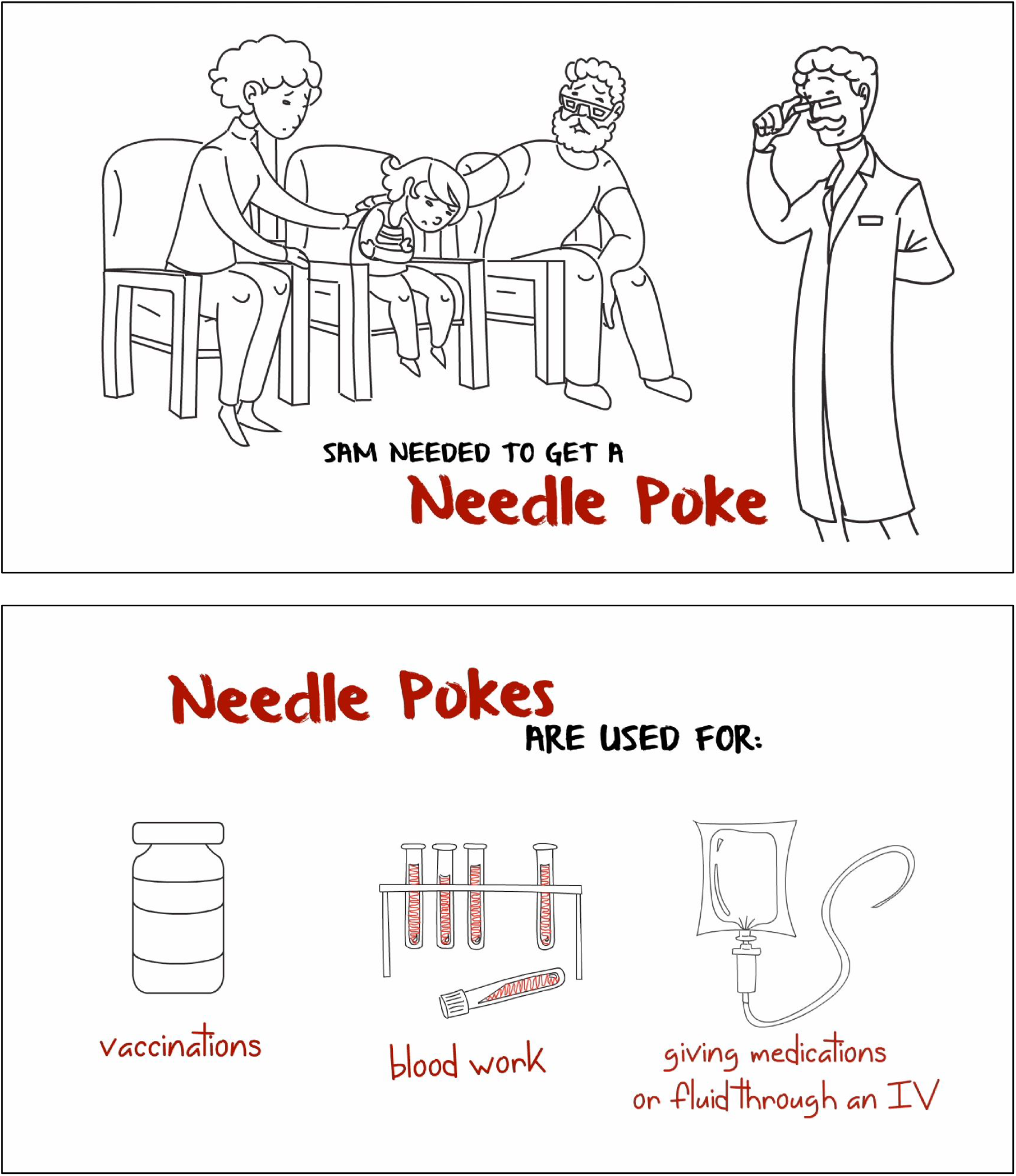
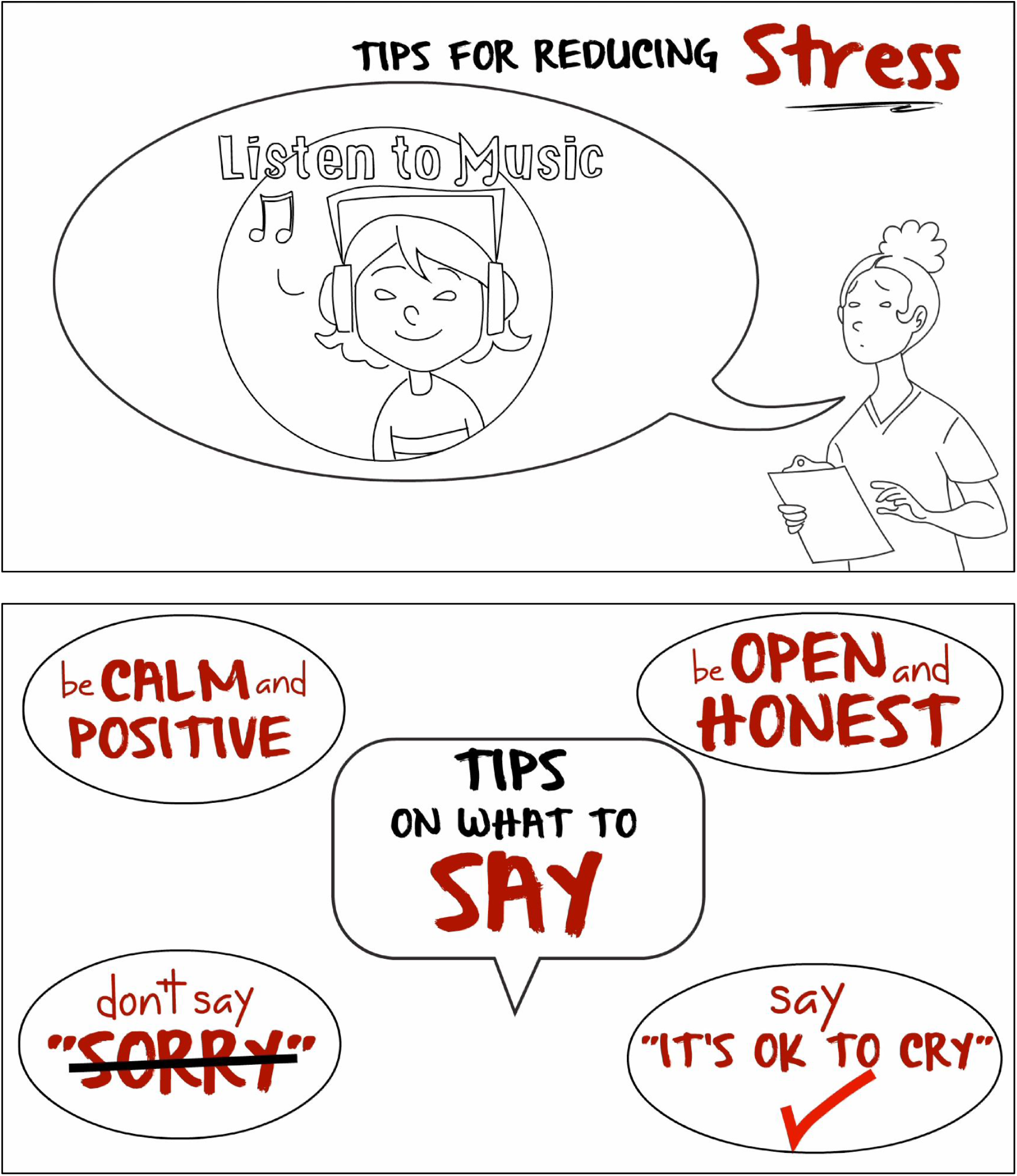
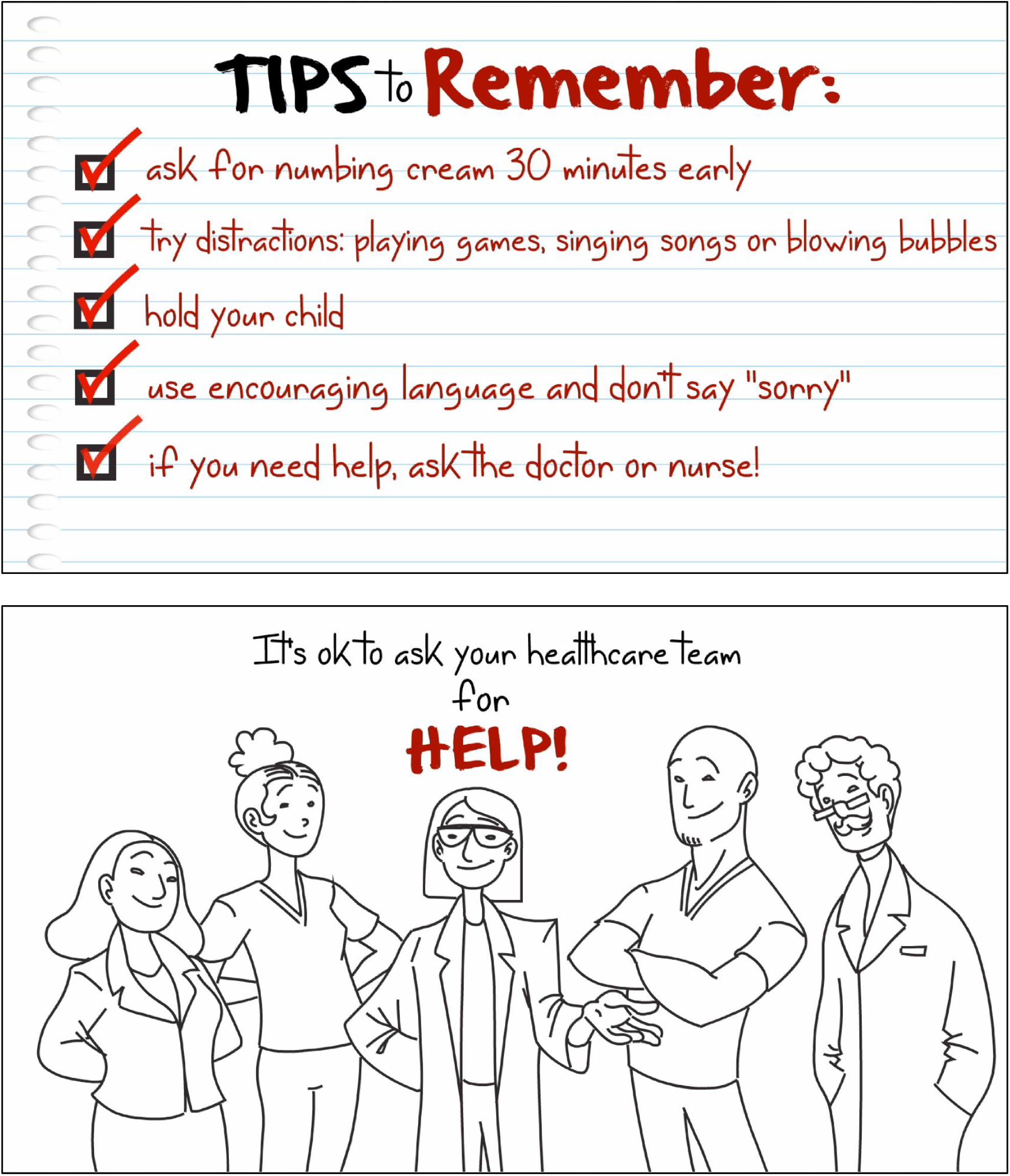

### Appendix C – Images from Usability Testing Infographic

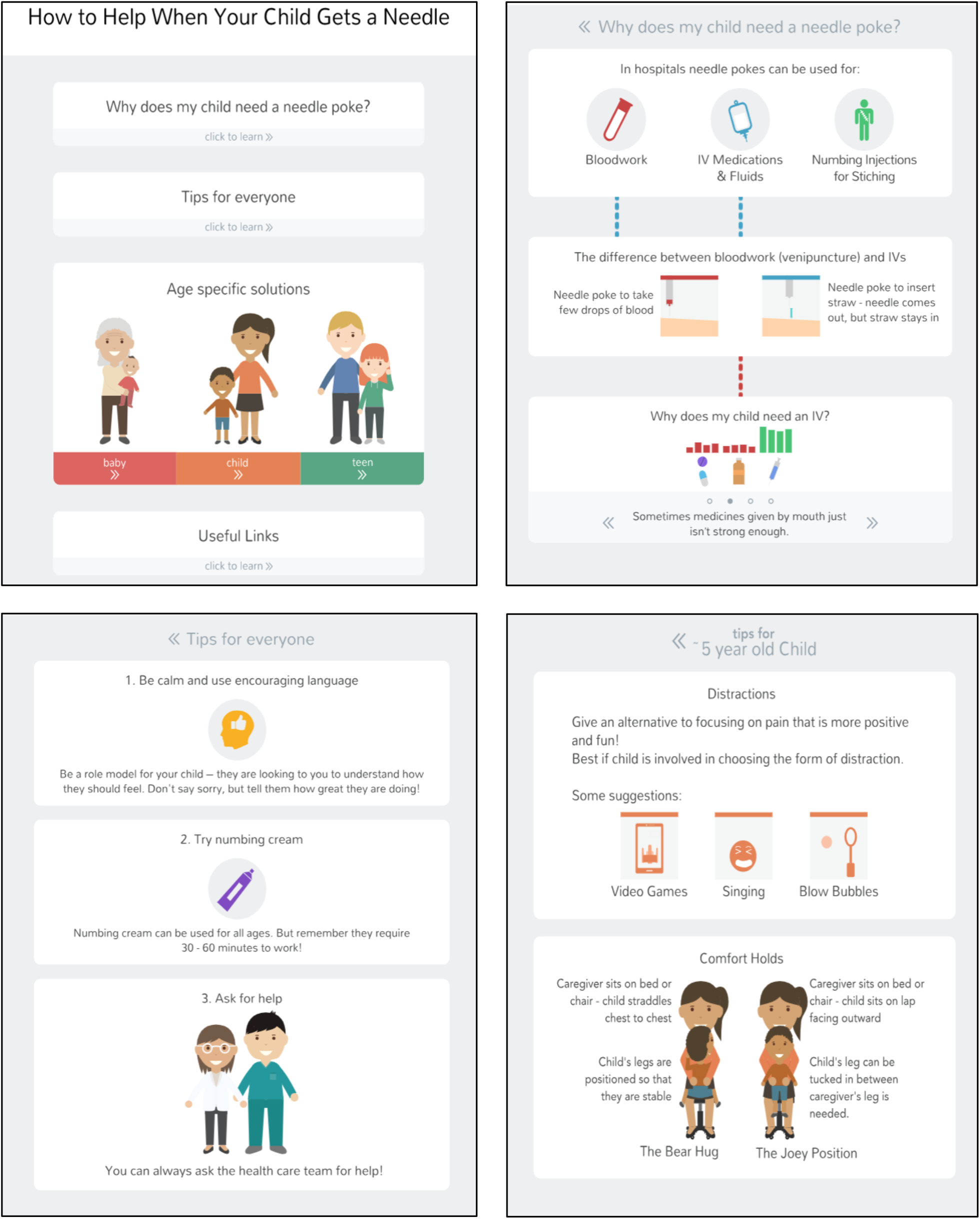

### Appendix D – Children’s Focus Group Interview Guide

Children (aged 7-14 years) and older children (15 years of age and older) will be interviewed within a focus group setting to understand their experience of procedural pain (such as needle insertion for IV) that took place in a hospital setting, including the language they use to describe aspects of the experience (e.g. “poke” vs. “needle”).

The focus group interview will be conducted with children in order to gain their perspective to help us define the concepts and language to use in information resources on pediatric procedural pain that we are developing for parents/families.

The focus group interview will open with this statement:

> *“We’re going to spend some time talking about when you got a needle poke at the hospital. We would like to know how you felt. We’re just going to talk about it. There are no needles here, this is not a hospital, and you will not be getting a poke today. If you start to feel sad or worried when we are talking, or if you just don’t feel like talking anymore, just say you want to stop or put your hand up, and we will stop talking, and you can go out to your [mom/dad/etc*.*]. You will still get to pick a toy*.*”*

#### The focus group interview will be guided by the following questions

1. What are some reasons that someone might have to get a needle poke?
2. Tell me about when you went to the hospital and got a poke. *Recall prompts:*
  a. Why did you go to the hospital? Why did you need the needle poke?
  b. Who were you with?
  c. What was the first thing that happened? What happened next?
  *(Use multiple pass approach to reflect back to children that they have said to see if more detail can be elicited)*
3. How did it make you feel? (A “Faces of Pain” scale may be used to help children express how they felt, e.g. children may be asked to identify or colour in the face that matches how they felt at different stages during the procedure.)
4. What kinds of other things did you see/hear/feel(touch)/smell etc. when you got the poke?
5. Did you or anyone else do anything to help it hurt less, or to help make it better for you? If “yes”, what was it? How did it make you feel? Did it help you? *Recall* prompts:
  a. Did you have anything to play with? (iPhone, iPad, etc.)
  b. Did you choose what to do? (i.e. keeping playing the video game)
  c. Was mom/dad/nurse talking to you or holding you?
  d. Did the nurse put cream on your arm?
6. What do you think might have helped make it better for you?? What would you tell the grown-ups to do to help you? Is there anything the grown-ups could say or do to make it easier for you? *(These questions will be asked last, which will help the children to re-establish their empowerment, after talking about the procedural pain experience in which they were disempowered*.)

**At the end of the focus group interview**, each child will choose a toy from a toy chest we provide. (Providing children the opportunity to choose will help re-establish their empowerment, after talking about the procedural pain experience in which they were disempowered.)

### Appendix E – Usability Survey

SECTION 1: Demographics

1) What is your gender?
  □ Male
  □ Female
3) What is your Age?
  □ Less than 20 years old
  □ 20-30 years
  □ 31-40 years
  □ 41-50 years
  □ 51 years and older
4) What is your Marital Status?
  □ Married
  □ Single
5) What is your gross annual household income?
  □ Less than $25,000
  □ $25,000-$49,999
  □ $50,000-$74,999
  □ $75,000-$99,999
  □ $100,000-$149,999
  □ $150,000 and over
6) What is your highest level of education?
  □ Some high school
  □ Some high school
  □ Some high school
  □ High school diploma
  □ Some post-secondary
  □ Post-secondary certificate/diploma
  □ Post-secondary degree
  □ Graduate degree
  □ Other
7) How many children do you have?——
8) How old are your children?————

SECTION 2: Assessment of attributes of the arts-based, digital tools

***\*participant is randomized to view 1 of 2 digital tools then automatically directed to the survey***

1. It is useful. [5-point Likert Scale]
2. It provides information that is relevant to me as a parent. [5-point Likert Scale]
3. It is simple to use. [5-point Likert Scale]
4. I can use it without written instructions or additional help. [5-point Likert Scale]
5. Its length is appropriate. [5-point Likert Scale]
6. It is aesthetically pleasing (i.e., images, colours, etc.). [5-point Likert Scale]
7. It helps me to make decisions about my child’s health. [5-point Likert Scale]
8. I would use it in the future. [5-point Likert Scale]
9. I would recommend it to a friend. [5-point Likert Scale]
10. List the most negative aspects: [open text]
11. List the most positive aspects: [open text]

### Appendix F – Images from Final Video

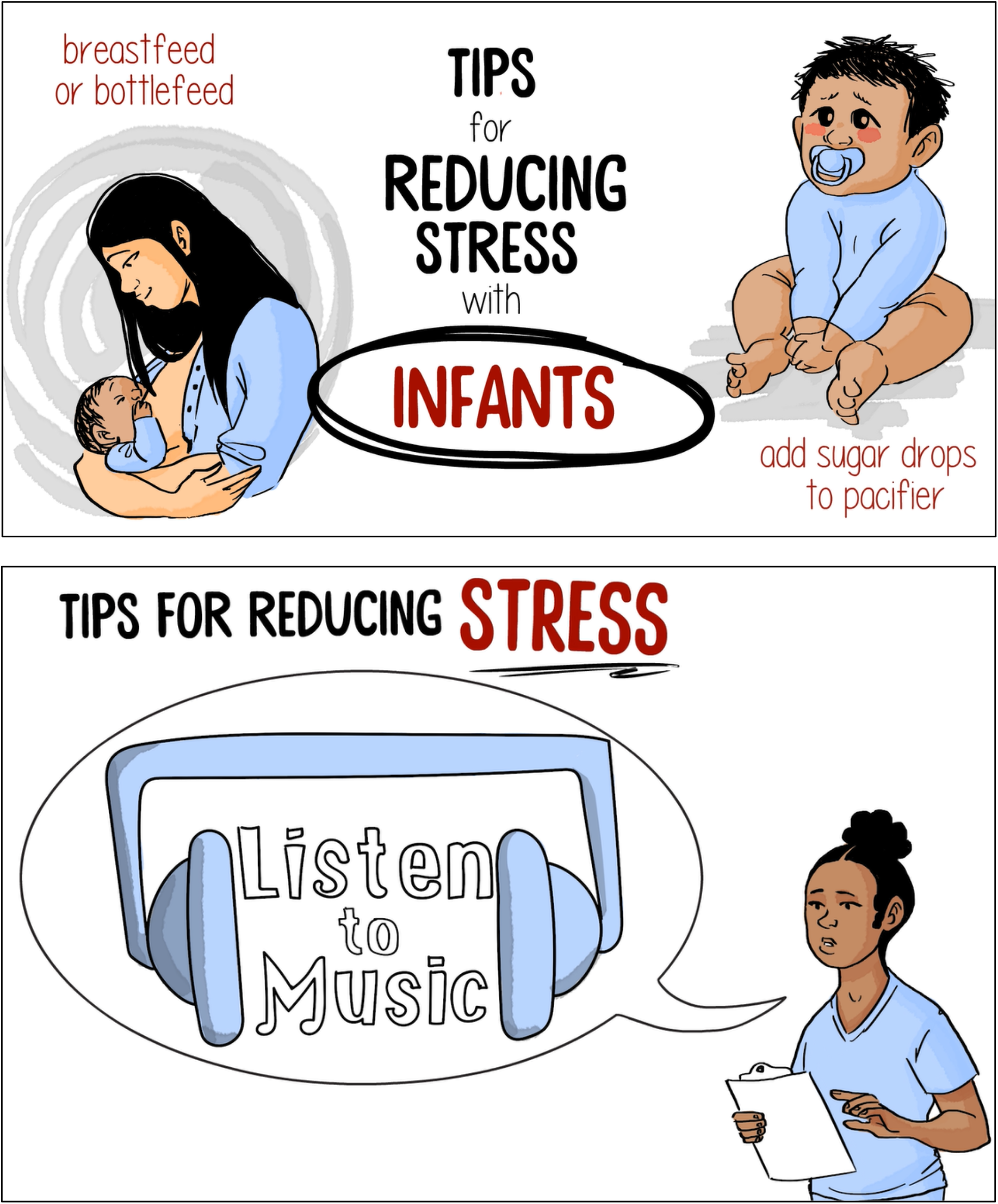
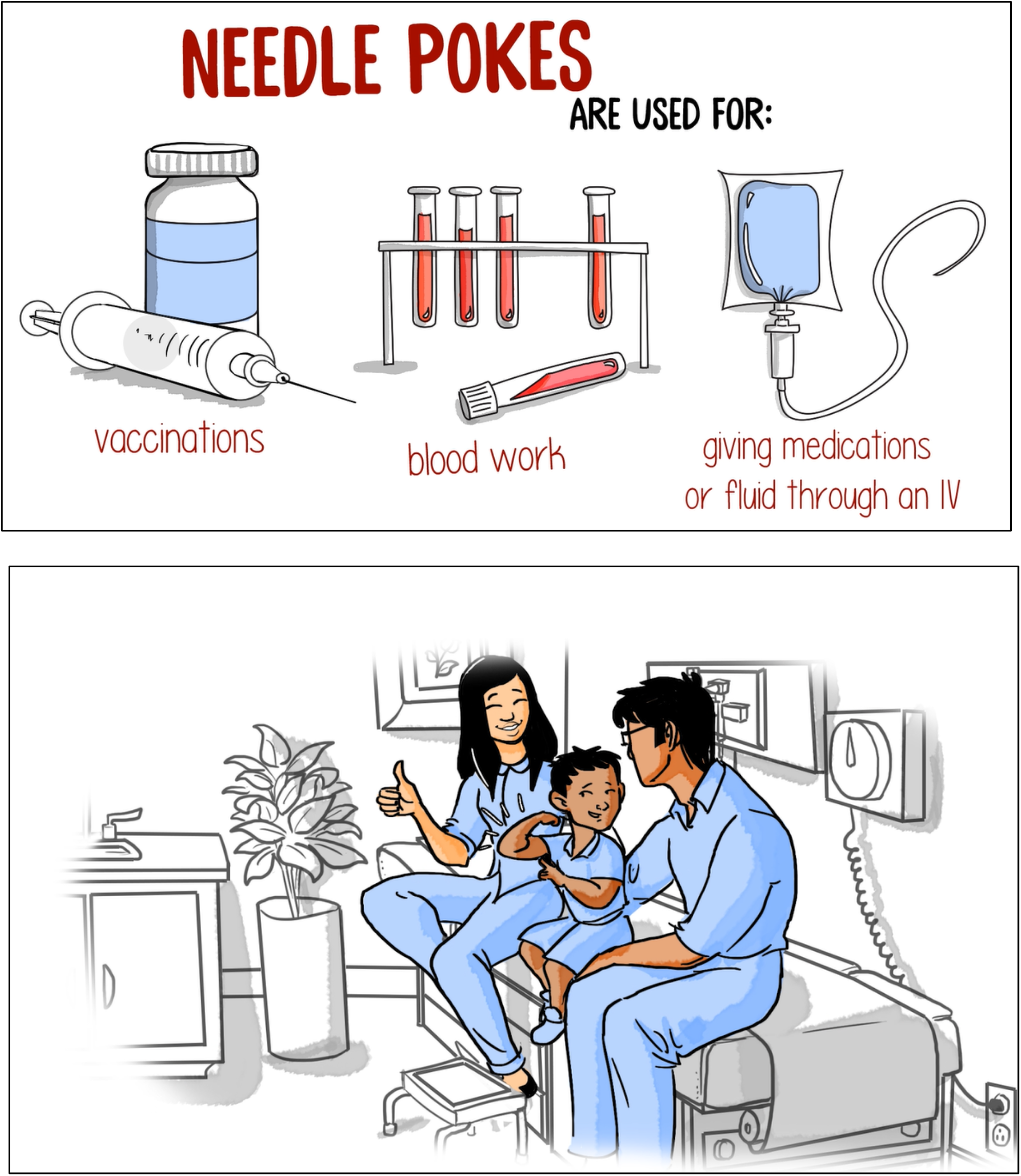

### Appendix G – Images from Final Infographic

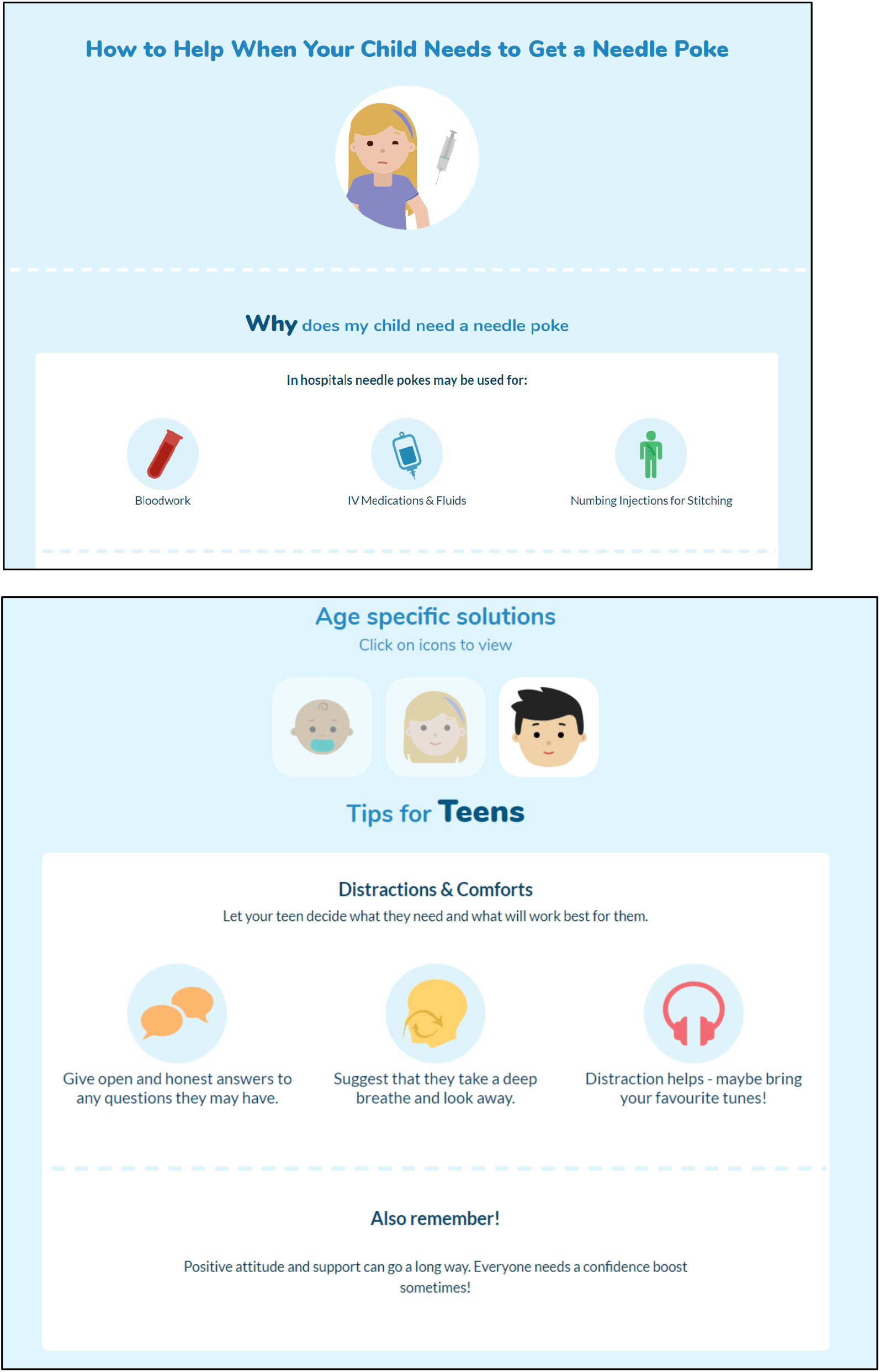
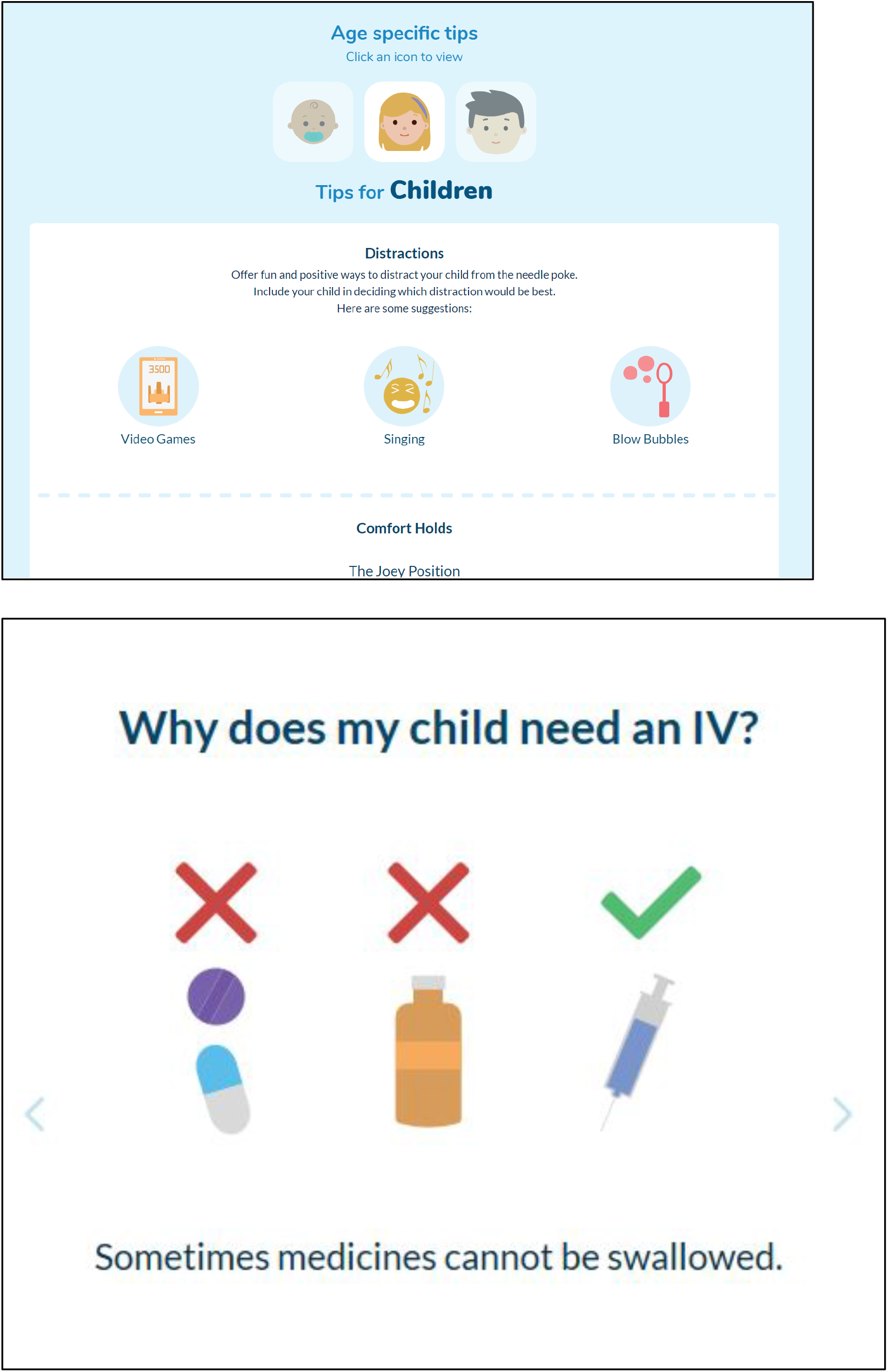

### Appendix H – Project Timeline

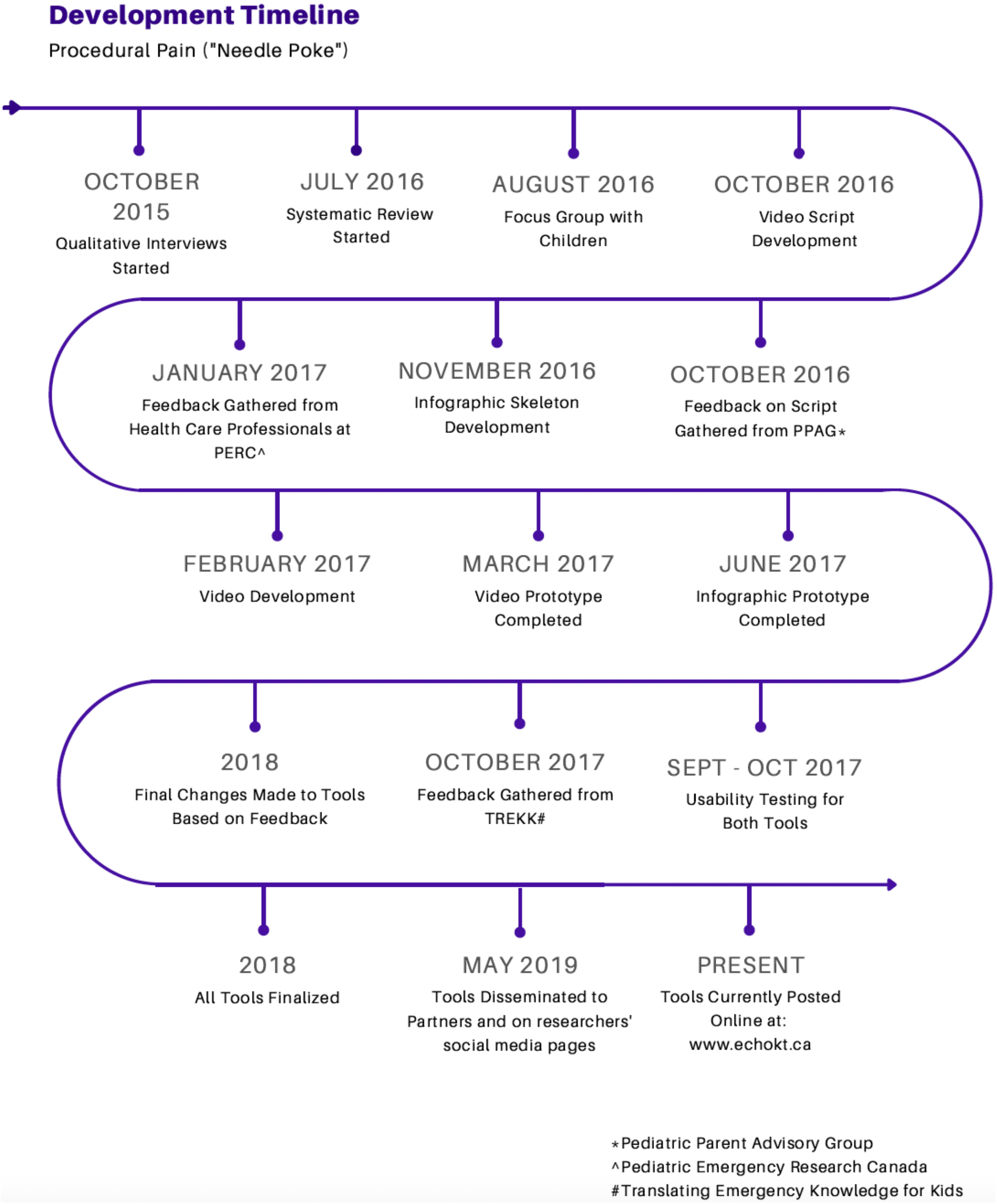

